# Obesity as a cause of kidney disease – insights from Mendelian randomisation studies

**DOI:** 10.1101/2020.09.13.20155234

**Authors:** Xiaoguang Xu, James M. Eales, Xiao Jiang, Eleanor Sanderson, David Scannali, Andrew P. Morris, Tomasz J. Guzik, Fadi J. Charchar, Michael V. Holmes, Maciej Tomaszewski

## Abstract

**Objective:** To examine if modifiable anthropometric indices of obesity exert putatively causal effects on different measures of kidney health and disease.

**Design:** Conventional observational and Mendelian randomisation study.

**Setting:** UK Biobank and international genome-wide association studies.

**Participants:** Approximately 300,000 participants of white-British ancestry from UK Biobank and up to 480,000 participants of predominantly European ancestry from genome-wide association studies.

**Main outcome measures:** Estimated glomerular filtration, blood urea nitrogen, kidney health index, chronic kidney disease, hypertensive renal disease, renal failure, acute renal failure, other disorders of kidney and ureters, IgA nephropathy and diabetic nephropathy.

**Results:** The Mendelian randomisation analysis indicated that increasing values of genetically predicted body mass index (BMI) and waist circumference were causally linked to changes in renal function indices including reduced estimated glomerular filtration and increased blood urea nitrogen in UK Biobank individuals. These associations were replicated using data from CKDGen Consortium individuals. One standard deviation increase in genetically-predicted BMI and waist circumference decreased the relative odds of kidney health index by 14% and 18% (OR=0·86; 95%CI: 0·82-0·92; P=9·18×10^-6^ for BMI and OR=0·82; 95%CI: 0·75-0·90; P=2·12×10^-5^ for waist circumference, respectively). Approximately 13-16% of the causal effect of obesity indices on kidney health was mediated by blood pressure. Obesity increased the risk of both acute and chronic kidney disease of several aetiologies including hypertensive renal disease and diabetic nephropathy.

**Conclusions:** These findings indicate that obesity is causally linked to indices of renal health and the risk of different kidney diseases. This evidence substantiates the value of weight loss as a strategy of preventing and/or counteracting a decline in kidney health as well as decreasing the risk of renal disease.

**What is already known on this topic:**

- Several previous studies reported associations between increasing adiposity/obesity and either a decline in kidney function or the increased risk of specific kidney disorders.
- High blood pressure and diabetes represent the two most common aetiologies of chronic kidney disease and co-exist with obesity.
- It is not clear to what extent these associations may reflect cause-effect relationships.

**What this study adds:**

- Our results show a consistent pattern of causation between obesity and kidney health and disease, across different biochemical parameters of kidney function and a broad spectrum of kidney health/disease.
- We further demonstrate that the causal effect of obesity on kidney health is partially mediated by blood pressure and largely independent of type 2 diabetes.
- We provide evidence for a causal relationship between two clinical indices of obesity and specific renal disorders including hypertensive renal disease, acute renal failure, chronic kidney disease and diabetic nephropathy.

## Introduction

Chronic kidney disease (CKD) affects more than 10% of adults worldwide and is predicted to become a global threat to public health.^1^ Interventions with reliable evidence for effectiveness in preventing CKD and/or slowing the progression of CKD are limited. Amongst the potentially safest to implement, cheapest to introduce and generally accepted by both patients and clinicians are health behaviours/lifestyle modifications such as weight loss. Such lifestyle modifications are usually recommended as first-line interventions in primary/secondary prevention of cardiovascular disease (CVD)^2^ and in the management of patients with CKD.^3^ However, it is not clear to what extent modifiable health behaviours are effective in improving clinical indices of kidney health i.e. in slowing the decline in estimated glomerular filtration rate (eGFR). For example, NICE guidelines recommend maintaining a healthy weight since it is a safe “healthy life” strategy rather than due to its nephro-protective effects.^3^

A number of previous investigations reported associations between increasing adiposity/obesity and a decline in kidney function or the increased risk of kidney diseases.^4–10^ However, it is not clear to what extent these associations reflect causality i.e. a cause and effect relationship between obesity and renal health/disease. This is of considerable relevance to clinical management given that only health behaviours with evidence of causal effects on disease (and/or its defining traits) are likely to succeed in effective prevention and treatment.^11^ In contrast, interventions targeting factors external to the disease-related causal pathways often fail to deliver the expected clinical outcomes.^12^

Mendelian randomisation (MR) has emerged as a genetic epidemiological approach that enables causal inference (analogous to a natural randomised controlled trial – RCT) and a powerful alternative and/or complement to the conventional RCT.^12^ Indeed, conducting RCTs can be very expensive, logistically challenging, risky (as the intervention may not demonstrate clinical efficacy), time-consuming and under certain circumstances – may not be free of clinical hazards.^13^ In contrast, MR does not require the resources or time consumed by RCT, is safe and robust.^14,15^ It relies on randomly assigned (at meiosis) genetic variants as proxies (or instruments) for an exposure (e.g. a modifiable lifestyle factor) to examine its putative causal effect on a clinical outcome. Compared to findings from observational studies, those from MR are immune to reverse causality and should be less susceptible to the influence of unmeasured confounding.^16,17^

MR-based analyses have rarely been applied to exposures of direct relevance to clinical nephrology despite their superiority over conventional observational studies.^18,19^ Previous studies used MR to uncover causality signals to or from renal phenotypes – i.e. an effect of eGFR on diastolic blood pressure (DBP), and risk of nephrolithiasis^20^ as well as the causal connection between higher blood pressure (BP) and the risk of microalbuminuria.^21^ Herein, using a set of MR studies, we sought to determine if obesity-defining traits exert putatively causal effects on a range of kidney phenotypes – from biochemical indices of kidney function to specific kidney diagnoses (Figure 1A).

**Figure 1.**
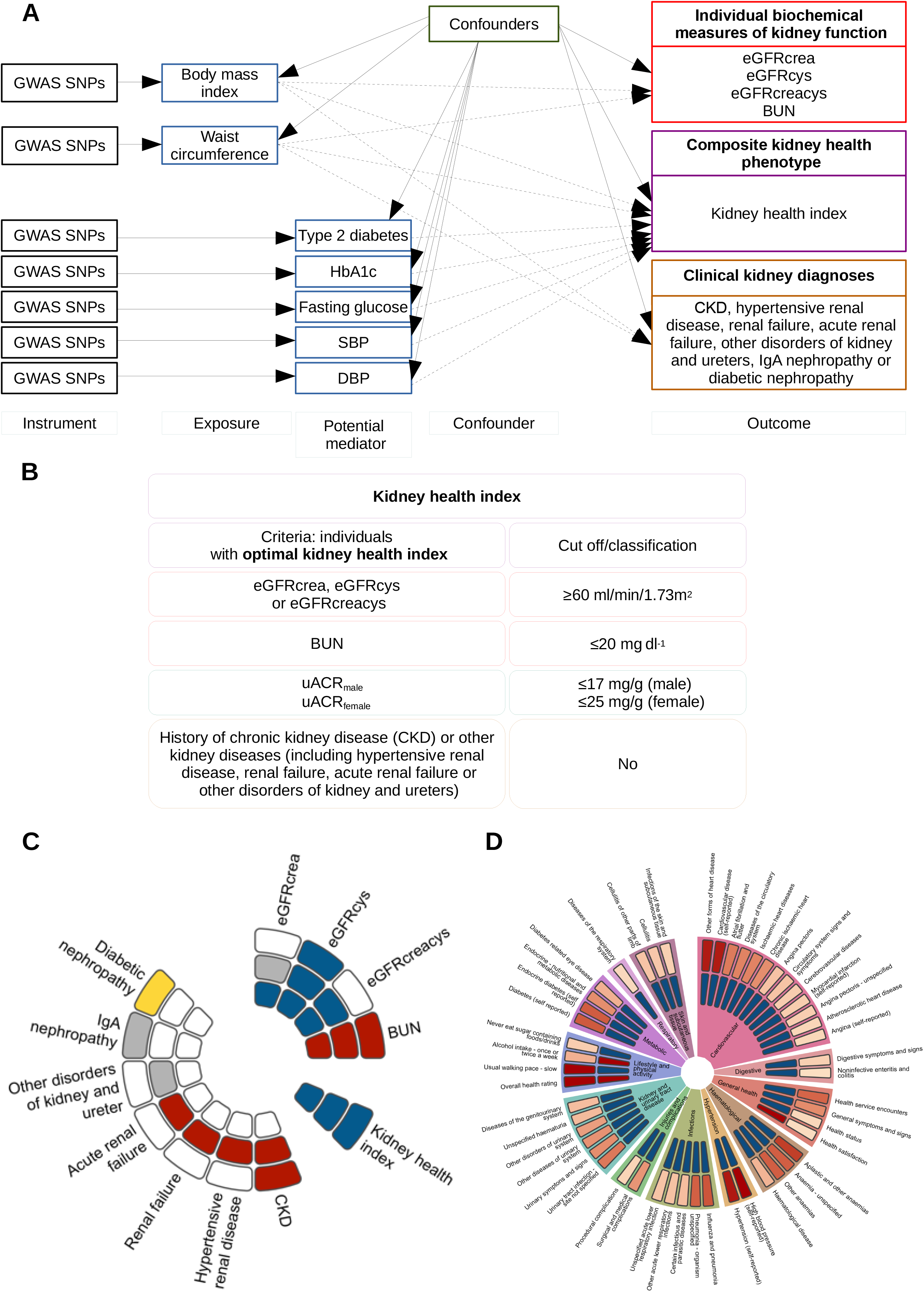
Overview of the strategy and the outputs of the study. **A**. Conceptual overview of the study showing hypothesised causal relationships (dashed line) between obesity indices (body mass index and waist circumference) and three different types of kidney phenotypes (i. individual biochemical measures of kidney function: eGFRcrea, eGFRcys, eGFRcreacys and BUN; ii. composite kidney health phenotype: kidney health index; iii. clinical kidney diagnoses (CKD, hypertensive renal disease, renal failure, acute renal failure, other disorders of kidney and ureters, IgA nephropathy and diabetic nephropathy) and hypothetical causal associations between diabetes/SBP/DBP and kidney health index. eGFRcrea – GFR estimated by creatinine, eGFRcys – GFR estimated by cystatin C, eGFRcreacys – GFR estimated by creatinine and cystatin C, BUN – blood urea nitrogen, SBP – systolic blood pressure, DBP – diastolic blood pressure. **B**. Criteria of kidney health index. **C**. Circular representation of information on associations between obesity indices (BMI and waist circumference) and kidney phenotypes. From outermost to innermost data circle we show: associations from two-sample MR, one-sample MR and observational analysis respectively. Associations were coloured as: white – not tested, grey – not associated with obesity indices, yellow – only associated with body mass index, red – positively associated with both obesity indices, blue – negatively associated with obesity indices. **D**. Circular representation of the 50 most highly correlated phenotypes with kidney health index. Strength of statistical significance is shown by colour in the outermost layer, ranging from pale red (least strongly associated) to dark red (most strongly associated), then the direction of correlation is shown, with positive values shown in red and negative values shown in blue. All phenotypes are grouped by clinical category which is labelled in the circle centre and demarcated by differing colours.

## Condensed methods

The overview of our strategy is shown in Figure 1A. Full details of the populations and methods are provided in supplementary material (Appendix).

### UK Biobank – key phenotypes

UK Biobank is a population-based resource on 487,395 individuals with a wide range of clinical data linked to genetic information. From this dataset we extracted information on the most common anthropometric measures of obesity (BMI and waist circumference) and four measures of kidney function (Figure 1A). In brief, we used information on blood urea nitrogen (BUN) and three measures of eGFR derived from: serum creatinine (eGFRcrea), cystatin C (eGFRcys) and both creatinine and cystatin C (eGFRcreacys). The number of individuals with informative values of eGFRcrea, eGFRcys, eGFRcreacys, BUN and was 304,800, 303,373, 317,425 and 314,731, respectively. We further generated the kidney health index – a novel composite renal phenotype integrating all available serum measures of kidney function (eGFRcrea, eGFRcys, eGFRcrea and BUN), a urinary bio-marker of kidney damage [albumin-to-creatinine ratio (uACR)] and the International Classification of Disease (ICD)-derived information on the history of kidney disease from Hospital Episodes Statistics (Figure 1B). A total of 217,289 individuals satisfied the criteria of optimal kidney health index and were defined as having optimal kidney health. The remaining 84,657 individuals did not meet at least one of the criteria of the kidney health index (and were defined as not having optimal kidney health).

### Observational analysis in UK Biobank

To characterise the observational association between directly measured BMI/waist circumference and quantitative serum bio-markers of kidney function (eGFRcrea, eGFRcys, eGFRcreacys, and BUN) we used linear regression. We applied logistic regression to examine the association between BMI/waist circumference and kidney health index. All these regression models were adjusted for age, age^2^ and sex, assessment centre and Townsend Deprivation Index.

We also explored the association between kidney health index (as an independent variable) and 403 binary traits (as a dependent variable) through logistic regression (with age, age^2^, sex, assessment centre and Townsend Deprivation Index as covariates). We selected all binary traits with a number of cases > 100 from self-reported data and ICD10-derived diagnoses available in UK Biobank (TableS1). These traits were further grouped into 22 clinical categories (TableS1). We calculated a correction for multiple testing using the false discovery rate (FDR) – findings with FDR<0·05 were considered statistically significant.

### Construction of genetic instruments

To build genetic instruments and generate genetic scores for BMI and waist circumference we followed the strategy from a previous study^22^. In brief, we used 72 and 43 instruments from SNPs associated with BMI and waist circumference, respectively, in GWAS conducted in 339,224 and 224,459 individuals by GIANT Consortium^23,24^ and validated in previous MR analysis^14^. To minimise the risk that potential causality signals from BMI/waist circumference to kidney phenotypes may reflect an effect on metabolism of creatinine/cystatin C/BUN rather than renal function, we further re-evaluated the associations by excluding SNPs within a distance of 500 Kbp of genes recognised for their roles in the metabolism of these blood biomarkers (TableS2). To examine if the effect of obesity on kidney-related traits was mediated by BP or diabetes, we further derived instruments/genetic scores for systolic blood pressure (SBP), DBP, type 2 diabetes, glycated haemoglobin (HbA1c) and fasting glucose. In the absence of summary statistics for BMI-unadjusted BP from previously published GWAS we applied a block jack-knife weighting approach^25^ to perform GWAS on SBP/DBP in UK Biobank. For type 2 diabetes, HbA1c and glucose, we used the respective summary statistics from previous GWAS^26–28^ that were not BMI-adjusted and independent of UK Biobank.

### Effect of metabolic parameters on kidney phenotypes – one-sample MR in UK Biobank

We first estimated the influence of obesity indices on kidney function measures in a one-sample MR using a two-stage least square approach (2SLS)^29^ with externally-derived genetic scores of BMI and waist circumference as an instrument, measured BMI and waist circumference as an exposure and kidney function parameters (i.e. eGFRcrea, eGFRcys, eGFRcreacys and BUN) as an outcome. The effect of BMI, waist circumference, SBP and DBP on kidney health index and/or ICD-informed kidney diagnoses (CKD, hypertensive renal disease, renal failure, acute renal failure or other disorders of kidney and ureters, where appropriate) (Figure 1A) was also tested using the 2SLS approach. Age, age^2^, sex, genotyping array, and first 10 genetic principal components were used as covariates in the above analyses. We calculated a correction for multiple testing at each experiment level using FDR – findings with FDR<0·05 were considered statistically significant.

### Effects of metabolic parameters on kidney phenotypes – two-sample Mendelian randomisation

We conducted two-sample MR using four different models (inverse variance weighted (IVW) regression, weighted median, RadialMR and MRPRESSO)^30–32^ to: (i) replicate the estimated causal effects of both obesity indices (as exposures) on biochemical parameters of kidney function; (ii) further investigate potentially causal effects of obesity on kidney diseases (CKD, IgA nephropathy and diabetic nephropathy) (Figure 1A); (iii) validate causal effects of BMI and waist circumference on kidney health index in a sensitivity analysis; (iv) explore the existence of reverse causality (i.e. causal effects from eGFRcys, BUN and kidney health index on obesity indices); (v) investigate whether diabetes and/or its defining traits (HbA1c and glucose) were causally related to kidney health index (Figure 1A). Summary statistics for exposures and outcomes and the selection of genetic instruments were derived from UK Biobank, CKDGen Consortium^33^ and DIAGRAM.^26^ To correct for multiple testing we calculated FDR at the experiment-wide level. Causal effect estimates from at least three of the four MR methods significant after the correction for multiple testing (FDR<0·05) were set as a criterion of evidence for causality.

### One-sample multivariable Mendelian randomisation analysis

The causal relationship between obesity indices, BP and the kidney health index in MR analyses motivated us to quantify whether the causal relationship from obesity indices to kidney health index were independent of BP. In brief, we conducted multivariable MR (MVMR) including obesity and BP as exposures and kidney health index as the outcome using a 2SLS approach.^34^ The results obtained from the MVMR estimation are the direct effect of each exposure on the outcome conditional on the other exposures included in the estimation. This was done separately for four combinations of exposures (BMI and SBP, waist circumference and SBP, BMI and DBP and waist circumference and DBP) with the genetic scores associated with both exposures included in the derivation of predicted estimates. The genetic scores were constructed using the same methods as in the MR above. Age, age^2^, sex, genotyping array, and first 10 genetic principal components were used as covariates. Full details of the MVMR analyses were described in Supplementary material (Appendix). We calculated a correction for multiple testing using FDR – findings with FDR<0·05 were considered statistically significant.

## Results

### Measures of general and abdominal obesity causally affect biochemical parameters of kidney function

We first used data from ≈300,000 unrelated UK Biobank individuals (TableS3) to examine how BMI and waist circumference relate to serum parameters of kidney function (eGFRcrea, eGFRcys, eGFRcreacys, BUN). Both directly measured obesity indices showed significant associations with each renal phenotype in the observational analysis, even after a correction for multiple testing (Figure 1C, Figure 2). Using genetically predicted information for BMI and waist circumference in one-sample MR, we detected causal relationships of both obesity measures with three indices of kidney function (eGFRcys, eGFRcreacys and BUN) (Figure 1C, Figure 2). The direction of these causal relationships was consistent – increasing obesity measures were related to reduced eGFR and increased BUN; this was apparent even after the correction for multiple testing (Figure 1C, Figure 2). We then sought to replicate the effect of BMI and waist circumference eGFRcys, eGFRcreacys and BUN in two-sample MR experiments. Of those, CKDGen Consortium derived summary statistics were available only for eGFRcys and BUN. We first replicated directionally consistent effect of BMI and waist circumference on both biochemical indices of kidney function in the IVW two-sample MR (TableS4, Figure 1C). Three additional MR models (weighted median, radial MR and MR-PRESSO) employed to minimise potential bias arising from horizontal pleiotropy^35^ confirmed these findings (TableS4). We then conducted sensitivity analyses using genetic scores constructed after exclusion of genetic variants mapping onto genes related to metabolism of creatinine/cystatin C/BUN (TableS2) – the association estimates between obesity measures and eGFRcys/BUN remained largely unaffected in one-sample MR (TableS5). Finally, we confirmed that there was no bidirectional causality between obesity measures and kidney function – neither BUN nor eGFRcys showed a causal effect on obesity measures (TableS6). Collectively, these results show a consistent and potentially causal contribution of higher BMI and waist circumference on the decline in kidney function across different MR models and in independent datasets.

**Figure 2.**
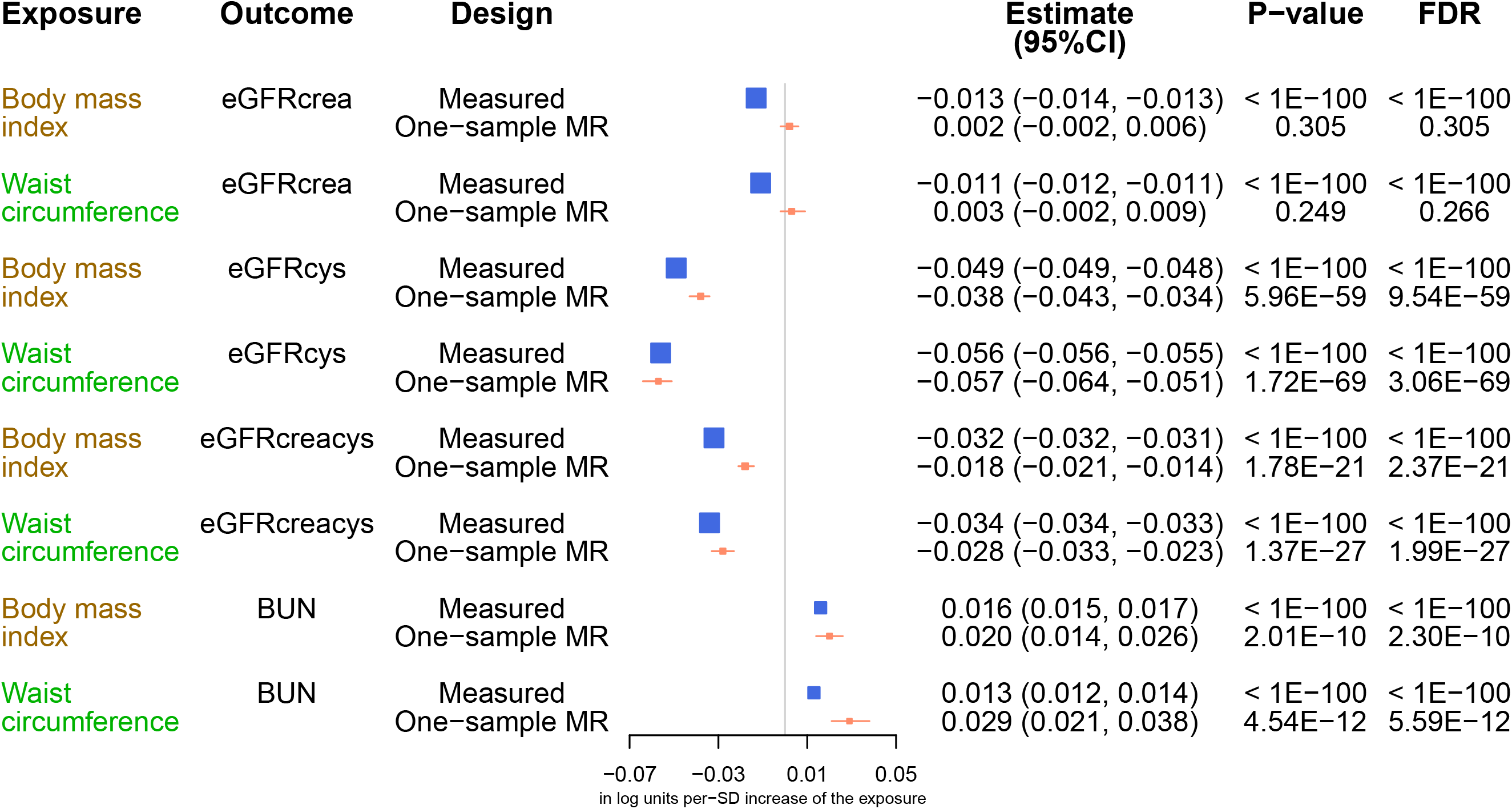
Relationships of BMI (brown) and waist circumference (green) on eGFR (eGFRcrea/eGFRcys/eGFRcreacys) and BUN from observational analyses and one-sample Mendelian randomisation analyses. Two different methods were coloured as: blue – observational analysis, orange – one-sample MR. MR – Mendelian randomisation, Estimate – effect size of each body weight measure on eGFR and BUN (in log units per-SD increase of the body weight measure), CI – confidence interval, P-value – level of statistical significance, FDR – false discovery rate.

### Obesity measures show an inverse causal relationship with kidney health index

Mindful of the potential limitation of using circulating concentrations of specific biomarkers as a marker of kidney function in MR,^36^ we generated a “kidney health index” as a composite binary phenotype derived from the blood and urine biochemistry combined with available clinical information in UK Biobank^37^ (see methods). As expected, those with optimal kidney health index showed statistically favourable renal profile across different serum bio-markers (eGFRcrea, eGFRcys, eGFRcreacys, BUN) and uACR when compared to the remaining individuals (TableS7). In observational analysis, both BMI and waist circumference were inversely related to the kidney health index (Figure 1C, TableS8). Further analysis of 403 binary traits in UK Biobank revealed strong associations between kidney health index and phenotypes expected to correlate with measures of kidney health and disease (kidney and urinary tract-related traits, cardiovascular disease, hypertension, diabetes and metabolic diseases) (Figure 1D, TableS9). Indeed, the directions of these associations were consistent with reduced odds of these disorders in those with optimal health index category. We then combined GIANT consortium-derived BMI and waist circumference genetic scores and the kidney health index as an outcome in one-sample MR. This analysis revealed that a one standard deviation (SD) genetically-predicted increase in BMI and waist circumference decreased the relative odds of kidney health index by 14% and 18% (OR=0·86; 95%CI: 0·82-0·92; P=9·18×10^-6^ for BMI and OR=0·82; 95%CI: 0·75-0·90; P=2·12×10^-5^ for waist circumference, respectively) (TableS8). We then used the genetic score of BMI and waist circumference excluding SNPs in/near genes involved in the metabolism of the blood biomarkers in one-sample MR; the association estimates between obesity measures and kidney health index remained unaffected (TableS5). Next, we employed two-sample MR tests with GIANT-derived genetic instruments for BMI and waist circumference (as exposures) and the summary statistics from our *de novo* GWAS of kidney health index (as an outcome) in UK Biobank. These sensitivity analyses confirmed the findings from one-sample MR (P_BMI_IVW_=2·27×10^-5^ and P_WC_IVW_=1·54×10^-4^) (TableS8, Figure 1C). Taken together, these results show that obesity measures show a potentially causal inverse effect on the overall kidney health index.

### The effect of obesity measures on kidney health index is not mediated by type 2 diabetes and largely independent of blood pressure

Elevated BP and diabetes have been proposed as the most likely biological mediators of the association between obesity and the risk of kidney disease.^38,39^ Therefore, we examined whether SBP, DBP, type 2 diabetes, HbA1c and fasting glucose are causally related to the kidney health index in UK Biobank (TableS10, TableS11). Of these, only SBP and DBP were causally related to the kidney health index in the expected direction – higher BP reduced the kidney health index (P_SBP_=6·26×10^-6^ and P_DBP_=1·69×10^-2^, respectively) (TableS10). Using one-sample multivariable Mendelian randomisation^34^ we then explored the extent to which the detected causality signals of BMI and waist circumference to kidney health index were mediated by BP. These analyses revealed that 13-16% of the causal effect of obesity indices on kidney health was mediated by BP (TableS8, TableS12-13). Collectively, these data show that potentially causal negative effect of obesity on kidney health is not mediated by type 2 diabetes and is largely independent of BP.

### BMI and waist circumference are causally related to increased risk of different kidney diseases

We then sought to investigate if BMI/waist circumference were causally related to clinically confirmed kidney outcomes using both one-sample and two-sample MR studies. Using GIANT Consortium-derived genetic scores for BMI and waist circumference and ICD10-derived diagnoses from Hospital Episodes Statistics we first conducted a series of one-sample MR in UK Biobank. We detected a causal relationship between measure of obesity and four out of five kidney diagnoses – increasing BMI and/or waist circumference increased the risk of hypertensive renal disease (OR=1·79; 95%CI: 1·14-2·82; P=1·15×10^-2^ for BMI and OR=2·41; 95%CI: 1·30-4·45 P=5·03×10^-3^ for waist circumference), renal failure (OR=1·51; 95%CI: 1·25-1·83; P=2·60×10^-5^ for BMI and OR=1·86; 95%CI: 1·43-2·42; P=4·16×10^-6^ for waist circumference) and CKD (OR=1·50; 95%CI: 1·16-1·96; P=2·44×10^-3^ for BMI and OR=1·83; 95%CI: 1·28-2·63; P=9·49×10^-4^ for waist circumference) (Figure 1C, TableS14). We then used two-sample MR using GWAS summary statistics of three kidney outcomes (CKD, IgA nephropathy and diabetic nephropathy) from populations independent to UK Biobank. These analyses replicated a causal effect of obesity on CKD (OR_ivw_=1·16; 95%CI: 1·04-1·30; P=6·07×10^-3^ for BMI and OR_ivw_=1·18; 95%CI: 1·003-1·393; P=4·66×10^-2^ for waist circumference) and uncovered a causal relationship between BMI and diabetic nephropathy (OR_ivw_=1·92; 95%CI: 1·44-2·54; P=6·86×10^-6^) (TableS15, Figure 1C). Collectively, these data show that obesity increases the risk of acute and chronic kidney disease of several aetiologies.

## Discussion

Currently there is no cure for a majority of patients with kidney diseases and the management strategies focus generally on symptoms, reducing the burden of complications of kidney disease rather than reversing the decline in renal function. Our knowledge of effective and safe strategies with potential to prevent the development of kidney disease in those with seemingly healthy kidneys is also limited. Indeed, while several health behaviours have been linked to the pathogenesis of CKD,^40^ robust evidence in support of their causal effects on eGFR, kidney health and the risk of kidney disease is hitherto missing. Therefore, we applied MR to examine whether two of the most commonly used clinical indices of obesity are causally related to a wide range of kidney phenotypes; from serum indices of kidney function and biochemistry-informed measure of kidney health to acute and chronic kidney disease and both primary and secondary nephropathies. Our results demonstrate a consistent signal of causality between obesity and kidney health and disease, irrespective of the type of MR experiment (one-sample or two-sample) or MR modalities applied and across different biochemical parameters of kidney function as well as a wide spectrum of kidney health/disease. We demonstrate that the causal relationship between indices of obesity and kidney health is not driven type 2 diabetes and is largely independent of BP. Finally, we provide evidence for causal association of BMI and/or waist circumference with specific renal disorders including hypertensive renal disease, CKD and diabetic nephropathy.

The association between BMI and/or waist circumference and different measures of kidney health and disease have been reported before in observational (both cross-sectional and longitudinal) studies – increasing obesity correlates with increased incidence of CKD,^4–7^ end-stage renal disease^8^ and a drop in eGFR calculated based on serum levels of creatinine^9^ or cystatin C.^5^ The results from our initial observational analysis of >300,000 individuals are fully consistent with these findings. However, due to inherent limitations of observational analyses (including confounding and reverse causality)^17^ these data cannot provide insights into causal contributions of obesity indices to eGFR and as such are not sufficient to inform i.e. effective therapeutic or preventive strategies. To this end, the MR-derived findings showing the effect of higher BMI and waist circumference on a decline in kidney function are an important piece of evidence in favour of a potential utility of interventions targeting BMI and/or waist circumference to improve eGFR in individuals largely unaffected by CKD (in whom the relevance of obesity to kidney function is possibly most apparent).^5,10^ Both routine engagement in moderate-intensity regular physical exercise and different dietary interventions have been shown to achieve clinically relevant changes in both waist circumference and BMI.^41^ In this context, our data suggest that non-pharmacological strategies promoting weight loss may have potentially nephro-protective effects.

Potential causality signals to eGFR measured using specific biochemical biomarkers have some interpretational limitations.^36^ For example, causal effects between obesity indices and eGFR may reflect the effects of BMI or waist circumference on the metabolism of specific biomarkers used to estimate kidney function (such as creatinine, cystatin C or BUN) rather than glomerular filtration *per se*.^36,42^ This is particularly relevant to creatinine and cystatin C given their association with body size, muscle mass and/or adiposity.^43,44^ To ensure that the findings from our MR studies with kidney function as an outcome were potentially robust to these limitations, we have undertaken a number of mitigation strategies. Firstly, we conducted MR studies on four different indices of kidney function (generated using three different serum biomarkers) as outcomes to minimise the effect of single blood metabolite on the results and conclusions. Secondly, we conducted sensitivity analyses by eliminating from the genetic instrument variants mapped to genes metabolically or mechanistically related to the abundance of BUN, creatinine and cystatin C. Finally, we carried out MR studies using a composite kidney phenotype that integrates information not only from all available serum biochemistry biomarkers but also - urine analysis and clinical records. The diversity of sources used to generate this composite phenotype makes our kidney health index not only much less dependent on metabolism of specific biomarkers but also possibly more reflective of genuine kidney health than individual parameters derived from single blood biomarkers. Indeed, our observational analysis on correlating kidney health index with over 400 traits in UK Biobank suggests that it may capture information from a wide range of renal phenotypes. Future studies are warranted to explore its potential diagnostic and predictive utility.

We examined type 2 diabetes, HbA1c, fasting glucose, SBP and DBP as potential mediators of the effect of obesity on kidney health given the established role of both diabetes and hypertension in the development of CKD.^45,46^ Indeed, both diabetes and hypertension showed strong association with the kidney health index in our observational analysis across a wide range of clinical phenotypes in UK Biobank. The causal association between higher BP on lower scores in kidney health index revealed by our analyses lends further support to the notion that poorly controlled hypertension is detrimental to the kidney function and structure.^47^ Some of the previously conducted MR studies showed that higher values of genetically predicted BP lead to increased risk of albuminuria^21^ and hypertensive renal disease^48^ but did not demonstrate a causal effect of BP on kidney function parameters.^36^ This may suggest that the detected signal of causality from BP to kidney health index is driven primarily by either of these phenotypes (given that they were used to define the composite renal outcome). Most importantly however, BP does not fully account for the causal effect of both obesity measures on the kidney health index. This in turn suggests that reduction in BMI/waist circumference (i.e. through weight loss) and the lowering of BP (i.e. through appropriate antihypertensive treatment) should translate into improvement in kidney health through shared and potentially different biological mechanisms/pathways. Further studies are required to identify the biological underpinnings of these findings and clarify the extent to which these effects could operate in an additive manner.

We appreciate that as in any Mendelian randomisation analysis, not all the assumptions pertaining to the validity of genetic instruments are actually testable, i.e. that the genetic instrument must be independent of potential confounders, and should only affect the outcome through the exposure rather than through alternative pathways (that is, through horizontal pleiotropy).^13,49^ However, we optimised our statistical pipeline by including a number of strategies to ensure the robustness of our genetic instruments, undertaking different validation experiments and/or sensitivity analyses. As a part of those analyses, we found for example no evidence of “bidirectional causality”^50^ – i.e. that kidney health or kidney disease is causal to changes in BMI or waist circumference.

In summary, our data from a set of statistically robust models embedded in the principles of causal inferences highlighted a casual inverse associations between two most common clinical indices of obesity and a range of kidney health and disease-related phenotypes. These findings suggest that obesity has a negative effect on kidney function and increases the risk of different kidney disorders across a spectrum of different aetiologies. Consequently, interventions targeting obesity have a potential to improve kidney health and reduce the risk of kidney disease at both individual and population level. We anticipate that the findings will help to stimulate further research and drive the development of public health policies to improve kidney health and prevent/treat kidney disease through encouraging weight loss.

## Data Availability

The data supporting the findings from these investigations are available within the article and the supplementary data files or are available upon reasonable request to the authors.

## Acknowledgments

This work was supported by British Heart Foundation project grants PG/17/35/33001 and PG/19/16/34270 and Kidney Research UK grant RP_017_20180302 to MT. Access to UK Biobank data has been through approved project (46114). MVH works in a unit that receives funding from the UK Medical Research Council and is supported by a British Heart Foundation Intermediate Clinical Research Fellowship (FS/18/23/33512) and the National Institute for Health Research Oxford Biomedical Research Centre. TJG is supported by the European Research Council [ERC and InflammaTENSION; ERC-CoG-726318] and the National Centre for Research and Development of Poland (ERA-Net-CVD PlaqueFIGHT).

## Disclosures

MVH has collaborated with Boehringer Ingelheim in research, and in adherence to the University of Oxford’s Clinical Trial Service Unit & Epidemiological Studies Unit (CSTU) staff policy, did not accept personal honoraria or other payments from pharmaceutical companies.

## Ethical approval

Each participating study obtained informed consent from the study participants and approval from its institutional review board.

## Data sharing

No additional data available.

## Transparency statement

The lead author affirms that the manuscript is an honest, accurate, and transparent account of the study being reported; that no important aspects of the study have been omitted; and that any discrepancies from the study as planned (and, if relevant, registered) have been explained.

## Notes

### Competing Interest Statement

The authors have declared no competing interest.

### Author Declarations

Only publicly available anonymised data sets were used. No approval was required.

## References

1 Eckardt K-U, Coresh J, Devuyst O, et al. Evolving importance of kidney disease: from subspecialty to global health burden. The Lancet 2013;382:158–69. doi:10.1016/S0140-6736(13)60439-0

2 Williams B, Mancia G, Spiering W, et al. 2018 ESC/ESH Guidelines for the management of arterial hypertensionThe Task Force for the management of arterial hypertension of the European Society of Cardiology (ESC) and the European Society of Hypertension (ESH). Eur Heart J 2018;39:3021–104. doi:10.1093/eurheartj/ehy339

3 Chronic kidney disease - NICE CKS. https://cks.nice.org.uk/chronic-kidney-disease!scenarioRecommendation:3 (accessed 21 May 2020).

4 Fox CS, Larson MG, Leip EP, et al. Predictors of new-onset kidney disease in a community-based population. JAMA 2004;291:844–50. doi:10.1001/jama.291.7.844

5 Madero M, Katz R, Murphy R, et al. Comparison between Different Measures of Body Fat with Kidney Function Decline and Incident CKD. Clin J Am Soc Nephrol 2017;12:893–903. doi:10.2215/CJN.07010716

6 Xu H, Kuja-Halkola R, Chen X, et al. Higher body mass index is associated with incident diabetes and chronic kidney disease independent of genetic confounding. Kidney Int 2019;95:1225–33. doi:10.1016/j.kint.2018.12.019

7 Kuma A, Uchino B, Ochiai Y, et al. Relationship between abdominal adiposity and incident chronic kidney disease in young- to middle-aged working men: a retrospective cohort study. Clin Exp Nephrol 2019;23:76–84. doi:10.1007/s10157-018-1606-y

8 Vivante A, Golan E, Tzur D, et al. Body mass index in 1.2 million adolescents and risk for end-stage renal disease. Arch Intern Med 2012;172:1644–50. doi:10.1001/2013.jamainternmed.85

9 Oh H, Quan SA, Jeong J-Y, et al. Waist Circumference, Not Body Mass Index, Is Associated with Renal Function Decline in Korean Population: Hallym Aging Study. PLoS ONE 2013;8. doi:10.1371/journal.pone.0059071

10 Chang AR, Grams ME, Ballew SH, et al. Adiposity and risk of decline in glomerular filtration rate: meta-analysis of individual participant data in a global consortium. BMJ 2019;364:k5301. doi:10.1136/bmj.k5301

11 Roberts JD. Thyroid Function and the Risk of Atrial Fibrillation: Exploring Potentially Causal Relationships Through Mendelian Randomization. JAMA Cardiol 2019;4:97–9. doi:10.1001/jamacardio.2018.4614

12 Cornish AJ, Tomlinson IPM, Houlston RS. Mendelian randomisation: A powerful and inexpensive method for identifying and excluding non-genetic risk factors for colorectal cancer. Mol Aspects Med 2019;69:41–7. doi:10.1016/j.mam.2019.01.002

13 Holmes MV, Ala-Korpela M, Davey Smith G. Mendelian randomization in cardiometabolic disease: challenges in evaluating causality. Nat Rev Cardiol 2017;14:577–90. doi:10.1038/nrcardio.2017.78

14 Holmes MV. Human genetics and drug development. N Engl J Med 2019; 380:1076–9. doi: 10.1056/NEJMe1901565

15 Holmes MV, Richardson TG, Ference BA, Davies N, Davey Smith G. Integrating genomics with biomarkers and therapeutic targets to invigorate cardiovascular drug development. Nat Rev Cardiol 2020; In Press.

16 Davey Smith G, Ebrahim S. ‘Mendelian randomization’: can genetic epidemiology contribute to understanding environmental determinants of disease? Int J Epidemiol 2003;32:1–22. doi:10.1093/ije/dyg070

17 Davies NM, Holmes MV, Davey Smith G. Reading Mendelian randomisation studies: a guide, glossary, and checklist for clinicians. BMJ 2018;362. doi:10.1136/bmj.k601

18 Holmes MV, Davey Smith G. Can Mendelian Randomization Shift into Reverse Gear? Clin Chem 2019;65:363–6. doi:10.1373/clinchem.2018.296806

19 Mohammadi-Shemirani P, Sjaarda J, Gerstein HC, et al. A Mendelian Randomization-Based Approach to Identify Early and Sensitive Diagnostic Biomarkers of Disease. Clin Chem 2019;65:427–36. doi:10.1373/clinchem.2018.291104

20 Morris AP, Le TH, Wu H, et al. Trans-ethnic kidney function association study reveals putative causal genes and effects on kidney-specific disease aetiologies. Nat Commun 2019;10:29. doi:10.1038/s41467-018-07867-7

21 Haas ME, Aragam KG, Emdin CA, et al. Genetic Association of Albuminuria with Cardiometabolic Disease and Blood Pressure. Am J Hum Genet 2018;103:461–73. doi:10.1016/j.ajhg.2018.08.004

22 Carreras-Torres R, Johansson M, Haycock PC, et al. Role of obesity in smoking behaviour: Mendelian randomisation study in UK Biobank. BMJ 2018;361. doi:10.1136/bmj.k1767

23 Locke AE, Kahali B, Berndt SI, et al. Genetic studies of body mass index yield new insights for obesity biology. Nature 2015;518:197–206. doi:10.1038/nature14177

24 Shungin D, Winkler TW, Croteau-Chonka DC, et al. New genetic loci link adipose and insulin biology to body fat distribution. Nature 2015;518:187–96. doi:10.1038/nature14132

25 Shao J, Wu CFJ. A General Theory for Jackknife Variance Estimation. Ann Stat 1989;17:1176–97. doi:10.1214/aos/1176347263

26 Scott RA, Scott LJ, Mägi R, et al. An Expanded Genome-Wide Association Study of Type 2 Diabetes in Europeans. Diabetes 2017;66:2888–902. doi:10.2337/db16-1253

27 Soranzo N, Sanna S, Wheeler E, et al. Common Variants at 10 Genomic Loci Influence Hemoglobin A1C Levels via Glycemic and Nonglycemic Pathways. Diabetes 2010;59:3229–39. doi:10.2337/db10-0502

28 Scott RA, Lagou V, Welch RP, et al. Large-scale association analyses identify new loci influencing glycemic traits and provide insight into the underlying biological pathways. Nat Genet 2012;44:991–1005. doi:10.1038/ng.2385

29 Burgess S, Small DS, Thompson SG. A review of instrumental variable estimators for Mendelian randomization. Stat Methods Med Res 2017;26:2333–55. doi:10.1177/0962280215597579

30 Burgess S, Bowden J, Dudbridge F, et al. Robust instrumental variable methods using multiple candidate instruments with application to Mendelian randomization. *ArXiv160603729 Stat* Published Online First: 12 June 2016. http://arxiv.org/abs/1606.03729 (accessed 8 Jan 2018).

31 Bowden J, Spiller W, Del Greco M F, et al. Improving the visualization, interpretation and analysis of two-sample summary data Mendelian randomization via the Radial plot and Radial regression. Int J Epidemiol 2018;47:1264–78. doi:10.1093/ije/dyy101

32 Verbanck M, Chen C-Y, Neale B, et al. Detection of widespread horizontal pleiotropy in causal relationships inferred from Mendelian randomization between complex traits and diseases. Nat Genet 2018;50:693–8. doi:10.1038/s41588-018-0099-7

33 Köttgen A, Pattaro C. The CKDGen Consortium: ten years of insights into the genetic basis of kidney function. Kidney Int 2020;97:236–42. doi:10.1016/j.kint.2019.10.027

34 Sanderson E, Davey Smith G, Windmeijer F, et al. An examination of multivariable Mendelian randomization in the single-sample and two-sample summary data settings. Int J Epidemiol 2019;48:713–27. doi:10.1093/ije/dyy262

35 Hemani G, Bowden J, Davey Smith G. Evaluating the potential role of pleiotropy in Mendelian randomization studies. Hum Mol Genet 2018;27:R195–208. doi:10.1093/hmg/ddy163

36 Yu Z, Coresh J, Qi G, et al. A bidirectional Mendelian randomization study supports causal effects of kidney function on blood pressure. Kidney Int Published Online First: 23 May 2020. doi:10.1016/j.kint.2020.04.044

37 Bycroft C, Freeman C, Petkova D, et al. The UK Biobank resource with deep phenotyping and genomic data. Nature 2018;562:203–9. doi:10.1038/s41586-018-0579-z

38 Stenvinkel P, Zoccali C, Ikizler TA. Obesity in CKD—What Should Nephrologists Know? J Am Soc Nephrol 2013;24:1727–36. doi:10.1681/ASN.2013040330

39 Hall JE, do Carmo JM, da Silva AA, et al. Obesity, kidney dysfunction and hypertension: mechanistic links. Nat Rev Nephrol 2019;15:367–85. doi:10.1038/s41581-019-0145-4

40 Francisco ALMD, Fresnedo GF, Palomar R, et al. The renal benefits of a healthy lifestyle. Kidney Int 2005;68:S2–6. doi:10.1111/j.1523-1755.2005.09901.x

41 Ross R, Neeland IJ, Yamashita S, et al. Waist circumference as a vital sign in clinical practice: a Consensus Statement from the IAS and ICCR Working Group on Visceral Obesity. Nat Rev Endocrinol 2020;16:177–89. doi:10.1038/s41574-019-0310-7

42 Tin A, Köttgen A. Genome-Wide Association Studies of CKD and Related Traits. Clin J Am Soc Nephrol Published Online First: 14 May 2020. doi:10.2215/CJN.00020120

43 Tsai C-W, Grams ME, Inker LA, et al. Cystatin C- and creatinine-based estimated glomerular filtration rate, vascular disease, and mortality in persons with diabetes in the U.S. Diabetes Care 2014;37:1002–8. doi:10.2337/dc13-1910

44 Tomaszewski M, Charchar FJ, Maric C, et al. Inverse associations between androgens and renal function: the Young Men Cardiovascular Association (YMCA) study. Am J Hypertens 2009;22:100–5. doi:10.1038/ajh.2008.307

45 Hall JE, Kuo JJ, da Silva AA, et al. Obesity-associated hypertension and kidney disease. Curr Opin Nephrol Hypertens 2003;12:195–200. doi:10.1097/00041552-200303000-00011

46 Chan JM, Rimm EB, Colditz GA, et al. Obesity, fat distribution, and weight gain as risk factors for clinical diabetes in men. Diabetes Care 1994;17:961–9. doi:10.2337/diacare.17.9.961

47 Bidani Anil K., Griffin Karen A. Pathophysiology of Hypertensive Renal Damage. Hypertension 2004;44:595–601. doi:10.1161/01.HYP.0000145180.38707.84

48 Eales, J.M. et al. Uncovering novel genetic mechanisms of hypertension through multi-omic analysis of the kidney. Nat Genet 2020; Accepted.

49 Sekula P, M FDG, Pattaro C, et al. Mendelian Randomization as an Approach to Assess Causality Using Observational Data. J Am Soc Nephrol 2016;27:3253–65. doi:10.1681/ASN.2016010098

50 Sattar N, Preiss D. Reverse Causality in Cardiovascular Epidemiological Research. Circulation 2017;135:2369–72. doi:10.1161/CIRCULATIONAHA.117.028307

